# Assessing the magnitude of and factors associated to demand for long acting reversible and permanent contraceptive methods among sexually active women in Kenya

**DOI:** 10.1101/2020.11.12.20230383

**Authors:** Michael W. Waithaka, Peter Gichangi, Mary Thiongo

**Affiliations:** International Centre for Reproductive Health Kenya, Mombasa, Kenya; Technical University of Mombasa, Mombasa, Kenya; Ghent University, Ghent, Belgium

**Author notes:** Corresponding author information: Michael W. Waithaka, Address: P.O. Box 91109 - 80103, Mombasa, Kenya.

## Abstract

**Introduction:** Though there has been an increase in use of long acting and permanent methods of contraception (LAPMs) globally over the last decade, the rates of unintended pregnancy still remain high. In Sub-Saharan African and especially in Kenya, while there has been increase in the uptake of LAPMs over the past years, there are limited recently available evidences on the real magnitude, recent trends and factors associated to demand for LAPMs among sexually active women in the reproductive age.

**Methods:** In this study, we used data from seven survey rounds (2014 to 2018) of a nationally representative Performance Monitoring and Accountability 2020 (PMA2020) household and female survey. PMA2020 survey is based on a multi-stage cluster sample design to draw a probability sample of households and eligible females. In this analysis, both descriptive and inferential statistics were used for data analysis and interpretations. Statistical significance was considered for p-values <0.05. STATA 16.0 software was used for all analyses.

**Results:** Among the 17,455 sexually active women in the reproductive age included in this study, 83.1% had demand for LAPMs. The total demand for LAPMs significantly decreased from 85.0% in 2014 to 81.0% in 2018. Age of the respondents, marital status, parity, rural/urban residence, county of residence and access to Family Planning/contraceptives information via media in the last 12 months were significantly associated with demand for LAPMs.

**Conclusion:** The total demand for LAPMs among sexually active women in Kenya is high, with the bigger share of the demand being unmet. Targeted programs to satisfy the existing demand of LAPMs and address the issue of unmet need for LAPMs should be put in place to ensure no woman is left behind. Similarly, programmers/advocates should utilize the locally available channels e.g. the media to generate new demand and overturn the decreasing demand for LAPMs.

**STRENGTHS AND LIMITATIONS OF THIS STUDY:**

- The nationally representative Performance Monitoring and Accountability 2020 (PMA2020) surveys are modelled on the insight from Demographic and Health Survey (DHS).
- PMA2020 uses standardized questionnaires (with questions extracted from DHS which have been validated and are extensively used in FP programs) to gather data about households and individual females that are comparable across program countries and consistent with existing national surveys.
- There are strengths to the PMA2020 survey which include inclusion of a large sample of women, use of data collectors without medical training and real-time data collection with sufficient controls for quality assurance.
- There are some limitations to consider when interpreting the data. Some outputs of this study utilized pooled data from seven-rounds of PMA2020 survey.

## INTRODUCTION

Though there have been concerted efforts in advocating for and improving access to modern methods of contraception, the global rates of unintended pregnancy still remain high. Unintended pregnancies and consequently unintended childbearing can lead to serious health, economic, and social consequences for women and their families.(1,2) Unintended childbearing is associated with a number of adverse maternal behaviors and child health outcomes such as delayed or inadequate prenatal care, lack of breastfeeding, preterm births as well as effect on children’s self-esteem. (3–7) Furthermore, many undesired pregnancies end in induced abortion.(8) In 2012, the worldwide rate of unintended pregnancy was estimated at 53 per 1,000 women aged 15–44.(9) Between 2010 and 2014, 44% (90% UI 42–48) of pregnancies and 23% (90% Uncertainty Interval (UI) 22–26) of births were unintended globally.(10) Sedgh et. al. 2014 reported that the rates of unintended pregnancy continued to decline in Africa and in the Latin America and Caribbean region between 2008 and 2012 as they had from 1995 to 2008. The pace of decline increased in Africa, from about 0.5 percent per year to about 2.0 percent per year.(9) In 2014, the demographic and health survey (KDHS) estimated that 35.7% of the births in Kenya were either unwanted (10.3%) or wanted later (25.4%). (11)

Access to essential modern contraceptive commodities remains a great concern to redress global inequity.(12) In addition, modern methods of contraception have a vital role in preventing unintended pregnancies (13), allowing people to attain their fertility desires as well as determine the spacing of pregnancies. Over the last decade, there has been an increase in use of modern contraceptives worldwide. Between 2000 and 2019 modern contraceptive prevalence among married women of reproductive age (MWRA) increased worldwide by 2.1 percentage points from 55.0% (95% UI 53.7%–56.3%) to 57.1% (95% UI 54.6%–59.5%).(14) Over the same period, the proportion of the need for family planning (FP) satisfied by modern methods among MWRA worldwide changed from 74.3% (95% UI 73.0%–75.4%) to 75.8% (95% UI 73.4%–78.0%).(14). Ahmed et. al. 2019 estimated that since 2013 the overall weighted average annual rate of change in modern contraceptive prevalence rates in all women across nine sub-Saharan Africa settings (Burkina Faso, Ethiopia, Ghana, Kenya, Uganda, DR Congo, Niger, and Nigeria) was 1·92 percentage points (95% CI 1·14 to 2·70).(15) Despite these developments unmet need for FP in Africa still remain high, with 22 million (95% UI 18–21 million) and 20 million (95% UI 18–22 million) women of Eastern Africa and Western Africa respectively estimated to have unmet need for modern methods in 2019.(14) This unmet need is driven by lack of information about contraceptive options and their use, unavailability of services, limited supplies, limited choice of methods, limited access to services, fear of social disapproval or a husband’s opposition, religious or cultural beliefs, concern for contraceptive side-effects or impacts on health, poor quality of available services; users’ and providers’ bias against some methods; and gender-based barriers to accessing services. (16–20)

While nearly one third (32 percent) of married women in Kenya expressed their desire to wait two years or more for their next birth and almost half (47%) of them did not want to have another child in 2014, about eighteen percent of them had an unmet need for FP services (9.2 percent in need of spacing and 8.3 percent in need of limiting).(11) With regards to FP use, KDHS 2014 estimated that about 58 percent of currently married women and about 65.4 percent of the sexually active unmarried women were using a method of contraception (Permanent methods, long-acting reversible methods, short-acting methods and traditional methods).(11) Short-acting methods include injectables (including depo-medroxyprogesterone acetate (DMPA) and norethisterone enanthate (NET-EN)), contraceptive pills, condoms, LAM, diaphragms, spermicidal agents and emergency contraception. Previous studies have estimated that the 1-year typical-use failure rates are 15% for condoms, 8% for oral contraceptive pills (OCPs), and 3% for injectables. (21,22) In addition, though contraceptive effectiveness requires a consistent and timely re-supply (every three months for DMPA and two months for NET-EN and in the case of OCPs adherence with daily dosing (22)), continuation of these short-acting methods can be poor, with studies reporting 12-month continuation of DMPA as low as 23-28%. (23,24) Permanent methods include female sterilization and vasectomy while long-acting reversible methods include intrauterine devices (IUDs) (effective for at least 12 years) and implants (effective from 3 to 7 years depending on type). Long-acting reversible methods when removed, return to fertility is prompt.(25) Long acting reversible and permanent methods (LAPMs) are the most effective (99% or greater) methods of contraception available and are very safe and convenient for protection against unintended pregnancy.(26) According to world health organization (WHO) eligibility criteria, almost all women are eligible for IUDs, implants and sterilization.(27)

Though the uptake of LAPMs in Kenya have increased over the past years (from 8.3% in 2008 to 16.5% (female sterilization (3.2%), IUDs (3.4%) and implant (9.9%)) in 2014 among the married women and 4.3% in 2008 to 9.8% (female sterilization (1.9%), IUDs (1.1%) and implant (6.8%) among unmarried sexually active women) (11,28,29), there are limited recently available evidences on the real magnitude, recent trends and factors associated to demand for LAPMs among sexually active women in the reproductive age. This study seek to fill in this evidence gap which might be essential in planning and promotion of LAPMs which effectively reduce visits to health facilities in the fragile health system in Kenya (30–34).

## METHODS

### Study design

This study utilize data drawn from seven survey rounds (2014 to 2018) of the Performance Monitoring and Accountability 2020 (PMA2020) female survey questionnaire in Kenya. PMA2020 survey platform was implemented to track the progress towards FP2020 goals. The nationally representative PMA2020 household and female survey were based on a multi-stage cluster sample design to draw a probability sample of households and eligible females with urban-rural and counties as strata. In Kenya, the survey was based on smart phone-assisted technology to collect and transmit data to update key FP indicators every six (2014 and 2015) to twelve (2016 to 2018) months. Full details of PMA2020 sampling and survey methodology has been published elsewhere.(35)

### Participants

In the PMA2020 survey, women aged 15–49 who are either usual members of the household or who slept in the household the night before the interview were eligible for the female interview. The female interview gathers information on sociodemographic characteristics and FP measures such as the current use of contraception, contraceptive use within 12 months preceding the interview among current non-users, reasons for not using or stopping a method of contraception among current non-users, intention to use contraception in the future among non-users, autonomy and influences related to contraceptive decision-making, and the contraception counseling at the health facility. In this analysis, we consider women in the reproductive age who are sexually active.

### Operational definitions

- **Sexually active women:** Women who were sexually active within the last four weeks preceding the survey.
- **Long acting reversible and permanent methods of contraception (LAPMs):** those methods that prevent pregnancy for three and more years per application (Implants, IUDs, male and female sterilizations).
- **Unmet need for long acting reversible FP methods:** A woman had unmet need for reversible long acting methods if she desired to delay or avoid pregnancy for 2 years or more but not using a FP method or is using unsuitable short acting method.
- **Unmet need for permanent FP methods:** A woman had unmet need for permanent FP methods if she had achieved the desired family size (don’t want any more children), but was not using LAPMs.

### Outcome variable

The outcome of interest in this study was total demand for LAPMs. Total demand for LAPMs was calculated as the sum of the sexually active who had unmet need for LAPMs and those who had their need for LAPMs met either using a longacting reversible contraception method or a permanent method of contraception. Sexually active women who desired to delay or avoid pregnancy for 2 years or more or did not want any more children (having a need for LAPMs) and were either not using any contraception, using traditional methods or were using short acting contraception methods were considered to have unmet need for LAPMs. On the other hand sexually active women with need for LAPMs and were either using long acting reversible or permanent contraception methods were considered to have their need for LAPMs met.

### Covariates

Consistent with extant literature on factors that influence demand for modern family planning, we assessed the following covariates:

- Socio-economic factors: Age, education level, wealth, marital status, rural/urban residence and county of residence
- Reproductive history factors: Parity, age at first sex and age at first birth
- Client related factors: Knowledge of modern contraceptives, knowledge of LAPMs, intendedness of the previous pregnancy, media exposure, source of contraceptives among users and who influence the decision to use among contraceptives users.
- Service related factors: Receipt of FP information from visiting provider or health care worker at the facility, counseled on side effects at the facility, counseled at the facility on what to do if they experienced side effects or problems and told of other FP methods

### Statistical analysis

Both descriptive and inferential statistics were used for data analysis and interpretations. Descriptive statistics were used to summarize the data. Bivariate analysis was conducted to assess the relationship between the covariates and the outcome variable. To estimate the effects of the covariates on demand for LAPMs, while controlling for possible confounders, a multivariable logistic regression model was fitted with factors that were significant in the bivariate analysis. Sampling weights, as well as clustering and stratification were taken into account in the analysis where appropriate. Statistical significance was considered for p-values <0.05. STATA 16.0 statistical software was used for all analyses (Stata Corporation, College Station, TX, USA).(36) In the analysis, sexually active women who were not sure if they want a (another) child, did not know how long they would like to wait till next birth and those who were pregnant at the time of the survey were excluded from the analysis.

### Ethical consideration

Ethical clearance for conducting PMA2020 study was obtained from the Kenyatta National Hospital/University of Nairobi ethical review committee (REF: P15/01/2014 and P801/09/2019), National Commission for Science, Technology and Innovation (REF: NACOSTI/P/14/0813/1676 and NACOSTI/P/19/2754). Interviews were conducted after informed consent from each eligible woman was obtained or parental consent was obtained as well as assent from the minors. All interviews were conducted in spaces which offered visual and audial privacy. The right of individual not to participate in the study was also respected.

## RESULTS

### Socio-demographic characteristics of respondents

As shown in Table 1, a total of the 17,455 sexually active women in the reproductive age were included in this study. Majority of the respondents were between the ages of 25–29 years (23.2%), had primary or technical level education (53.8%) and resided in rural areas (65.1%). Concerning marital status, 85.3% of respondents were married or in a union. About 62.5% of the respondents came from households in the lowest to middle wealth quintiles. About 13.1% of the respondents came from Nairobi County.

**Table 1:** Socio-demographic characteristics of respondents

| Variable | Unweighted<br>N | Weighted<br>N | Weighted<br>percent |
| --- | --- | --- | --- |
| Total | 17455 | 17455 | 100 |
| Age |  |  |  |
| 15-19 | 1053 | 1044 | 6.0 |
| 20-24 | 3268 | 3336 | 19.1 |
| 25-29 | 3940 | 4052 | 23.2 |
| 30-34 | 3311 | 3351 | 19.2 |
| 35-39 | 2497 | 2372 | 13.6 |
| 40-44 | 2003 | 1968 | 11.3 |
| 45-49 | 1383 | 1333 | 7.6 |
| Education |  |  |  |
| Never | 872 | 752 | 4.3 |
| Primary/Technical | 9437 | 9387 | 53.8 |
| Secondary | 4765 | 4862 | 27.9 |
| Higher | 2379 | 2453 | 14.1 |
| Missing | 2 | 1 | 0.0 |
| Wealth |  |  |  |
| Lowest quintile | 3888 | 3660 | 21.0 |
| Lower quintile | 3786 | 3753 | 21.5 |
| Middle quintile | 3493 | 3489 | 20.0 |
| Higher quintile | 3287 | 3217 | 18.4 |
| Highest quintile | 3001 | 3337 | 19.1 |
| Marital status |  |  |  |
| Never Married | 1947 | 1981 | 11.4 |
| Married/In a union | 14924 | 14891 | 85.3 |
| Widowed/Divorced | 576 | 576 | 3.3 |
| Missing | 8 | 8 | 0.0 |
| Residence |  |  |  |
| Urban | 6439 | 6099 | 34.9 |
| Rural | 11016 | 11356 | 65.1 |
| County |  |  |  |
| BUNGOMA | 2037 | 1606 | 9.2 |
| KAKAMEGA | 729 | 1507 | 8.6 |
| KERICHO | 1885 | 2037 | 11.7 |
| KIAMBU | 1562 | 1487 | 8.5 |
| KILIFI | 1746 | 1651 | 9.5 |
| KITUI | 1872 | 1742 | 10.0 |
| NAIROBI | 1501 | 2287 | 13.1 |
| NANDI | 2045 | 1756 | 10.1 |
| NYAMIRA | 1824 | 1357 | 7.8 |
| SIAYA | 1507 | 1511 | 8.7 |
| WEST POKOT | 747 | 515 | 3.0 |

### Calculating demand for LAPMs

As shown in Figure 1, out of the total number of sexually active women in the reproductive age, 1,304 (6.4%) were pregnant at the time of the survey, 1,457 (7.1%) women were not sure if they want a (another) child while 270 (1.3%) either did not know how long they would like to wait for next birth or had the wait time till next birth missing. These women were excluded from further analysis in this study. Of the 17,455 sexually active considered for analysis in this study, 3,061 (16.9%) respondents desired to have a child then or within 2 years or were infecund, menopausal or infertile. These women were considered not to have a need for LAPMs. An additional 6,651 (39.0%) women desired to delay or avoid pregnancy for 2 years or more while 7,743 (44.1%) women did not want any more children. These two groups of women were considered to have a need long acting reversible and permanent contraception.

**Figure 1:**
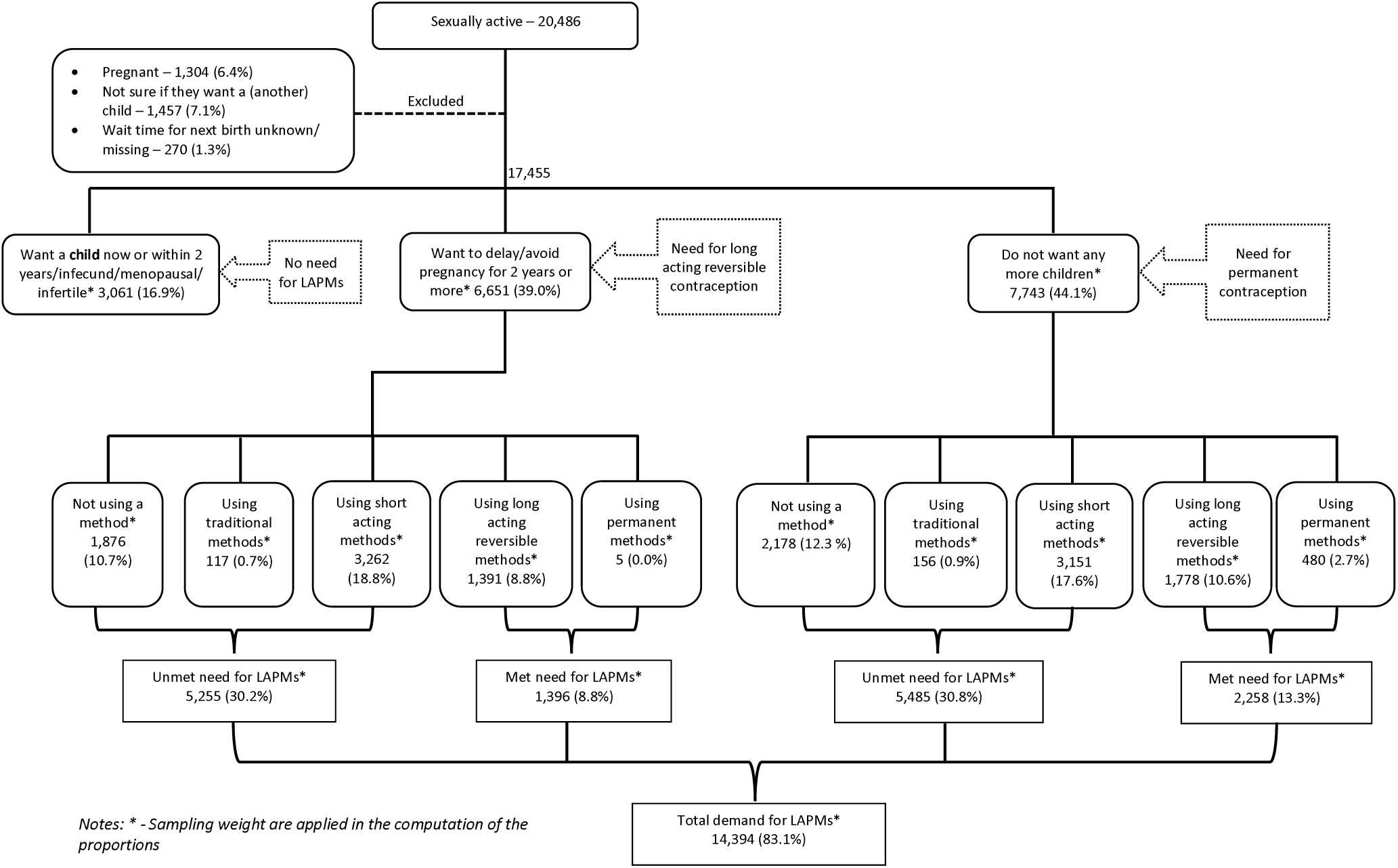
Calculating unmet need, met need and total demand for LAPMs among sexually active women

Of the women who desired to delay or avoid pregnancy for 2 years or more, 5,255 (30.2%) were either not using a contraception method (1,876 (10.7%)), using traditional methods (117 (0.7%)) or were using short acting contraception methods (3,262 (18.8%)) – and were considered to have unmet need for LAPMs. On the other hand about 1,396 (8.8%) of these women had their need for LAPMs met – either using long acting reversible or permanent contraception methods. Similarly among sexually active women who did not want any more children, 5,485 (30.8%) had unmet need for LAPMs (2,178 (12.3 %) not using a contraception method, 156 (0.9%) using traditional methods and 3,151 (17.6%) using short acting methods) while 2,258 (13.3%) had their need for LAMPs met (1,778 (10.6%) using long acting reversible methods and 480 (2.7%) using permanent methods).

About 14,394 (83.1%) sexually active women in the reproductive age were estimated to have demand for LAPMs (Figure 1).

### Trends in modern contraceptives use, demand and unmet need for LAPMs

Figure 3 shows trends in modern contraceptives use among sexually active women in Kenya between 2014 and 2018. The modern contraceptives use rate has significantly increased from 61.4% in 2014 to 67.4% in 2018 (p<0.001). We also note that as the rate in modern contraceptives use increase, the proportion of sexually active women using LAPMs significantly increase from 17.3% in 2014 to 30.3% in 2018 (p<0.001). In contrast, the proportion of sexually active women using short acting modern contraceptives significantly decrease from 44.1% in 2014 to 37.1% in 2018 (p<0.001).

**Figure 2:**
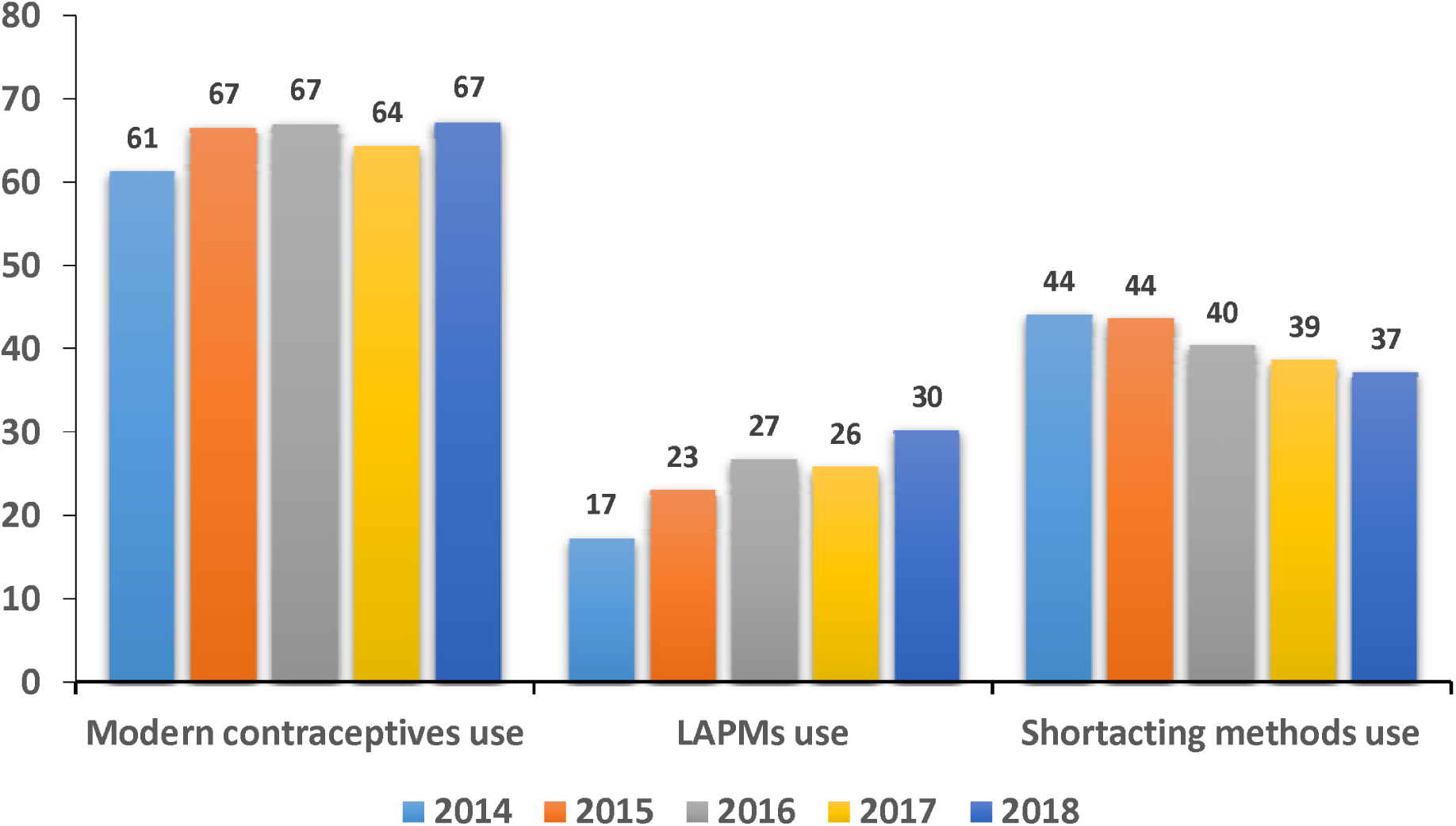
Trends in modern contraceptives use

**Figure 3:**
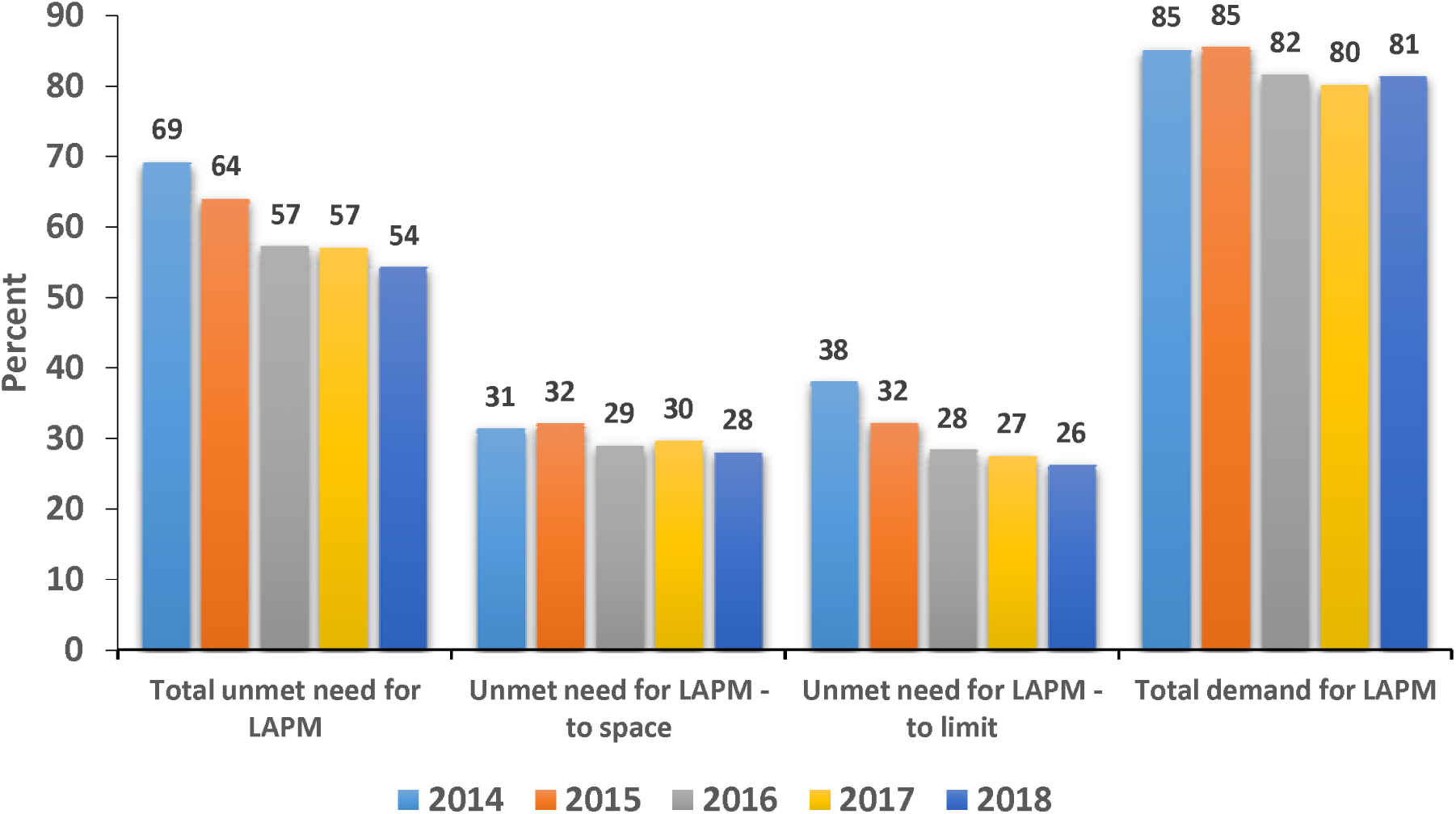
Trends in demand and unmet need for LAPM

Total demand for LAPMs and its components for sexually active women in Kenya between 2014 and 2018 is shown in Figure 3. From the graph, we observe that the total demand for LAPMs has significantly decreased from 85% in 2014 to 81% in 2018 (p<0.001). The percentage of sexually active women with unmet need for LAPMs has significantly decreased from 69.2% in 2014 to 54.2% in 2018 (p<0.001), about 15 percentage points. Almost all this decrease in unmet need was attributed to a decrease of unmet need for limiting, which dropped from 38.0% in 2014 to 26.2% in 2018. Unmet need for spacing dropped slightly from 31.2% in 2014 to 28.0% in 2018.

### Factors associated with demand for LAPMs

Results in Table 2 shows high demand for LAPMs across all the categories of all covariates (>75%) apart from among sexually active women who did not have any children (64.8%), sexually active women from West Pokot (54.3%) and sexually active women with no formal education (67.4%). Bivariate analyses (Table 2) indicated there was no evidence of an association between either, knowledge of any FP method, knowledge of LAPMs, who made the final decision to use the method, counseling on side effects when the women obtained the current method or being told about other FP method options the women could use and demand for LAPMs. There was however an association between demand for LAPMs and a participant’s age, marital status, household wealth, education level, parity, rural/urban residence, county of residence, access to FP information via media in the last 12 months, receipt of FP information from health care workers, source of the current method of contraception, intendedness of the last pregnancy and counseling on what to do if the respondent experienced side effects. The greatest predictor of demand for LAPMs among the sexually active women was the number of children the participant had (parity).

**Table 2:**
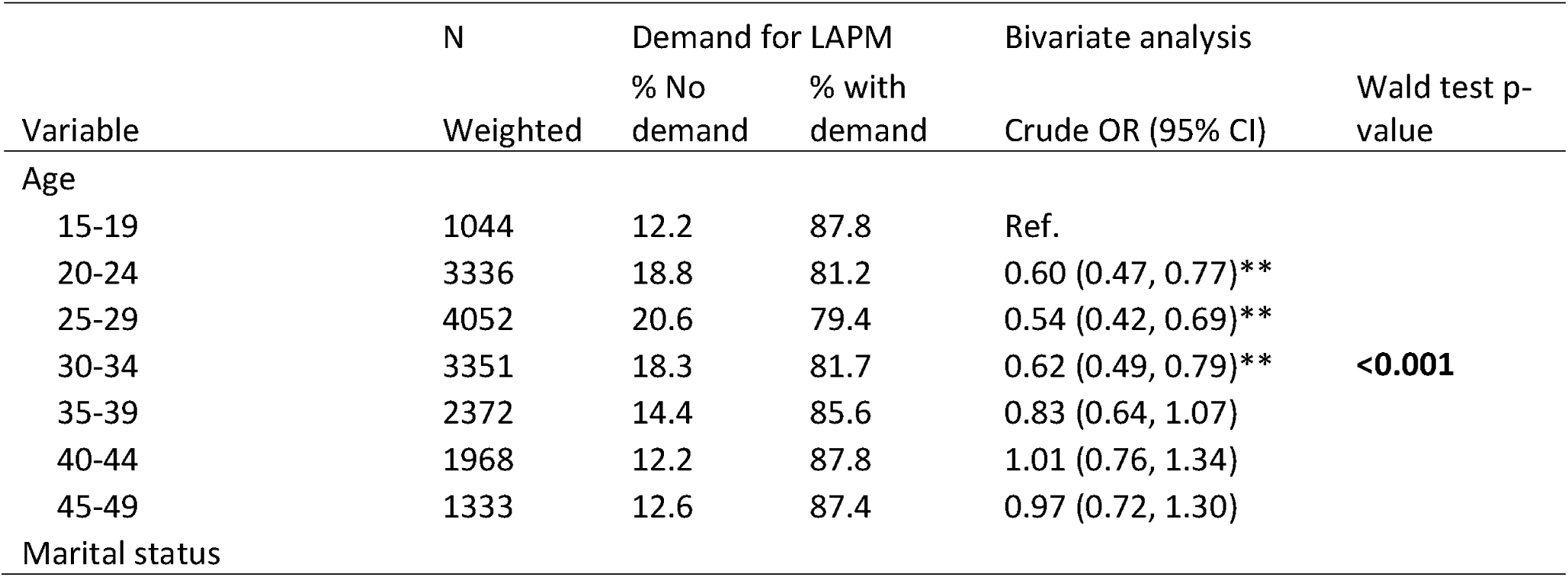
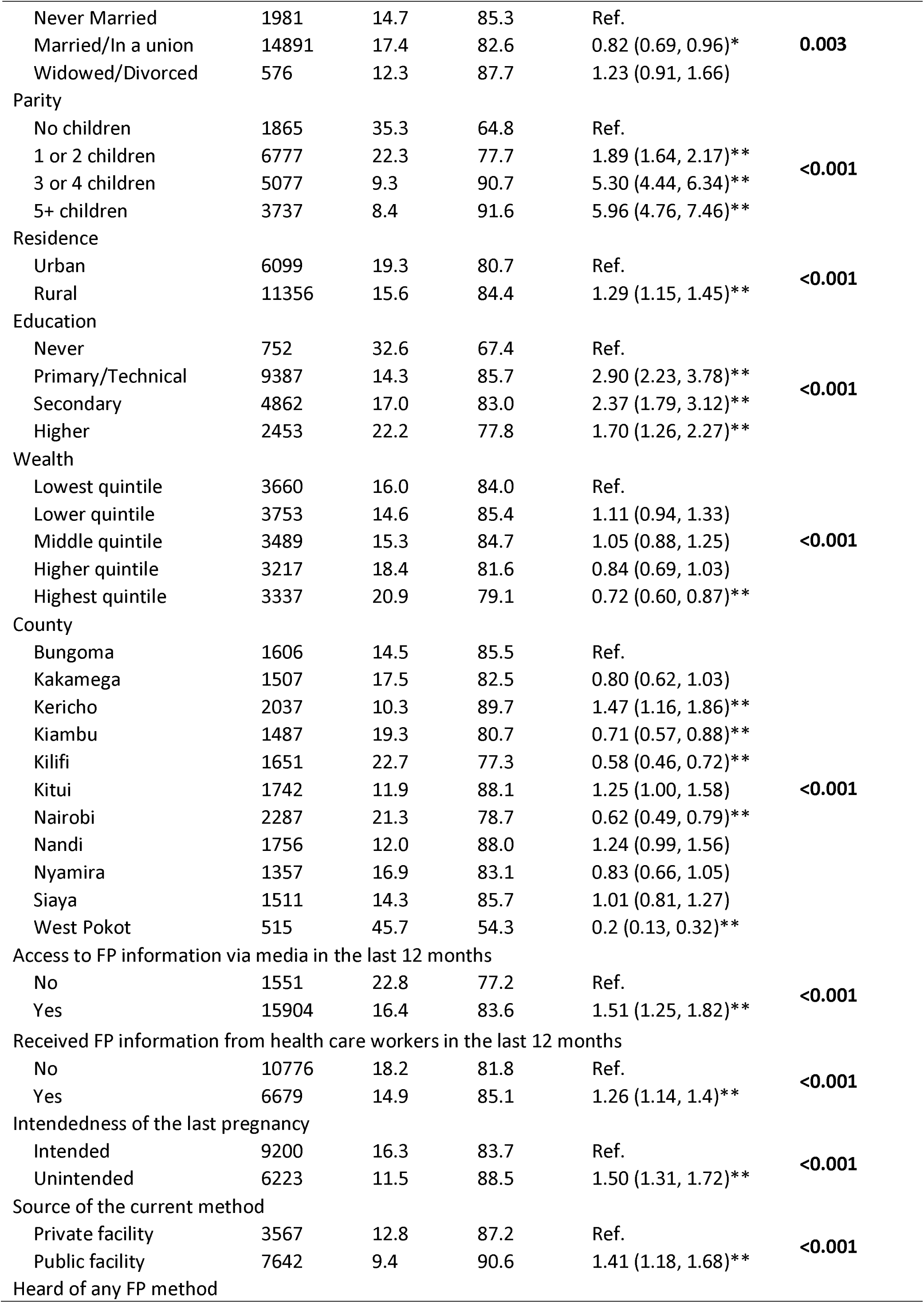
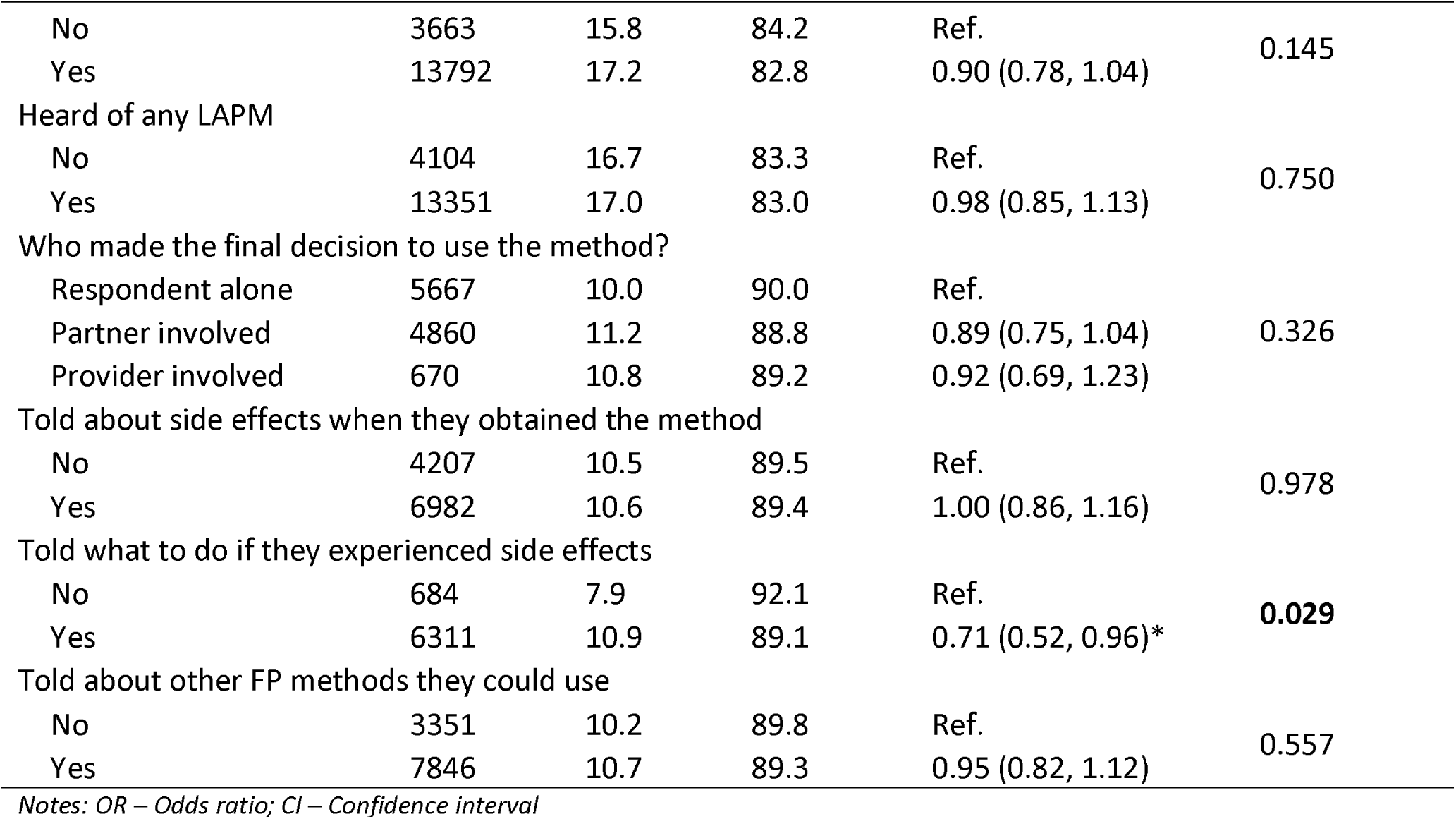
Bivariate analysis to identify respondents’ characteristics that may be associated with demand for LAPMs

To identify variables associated with the demand for LAPMs after adjusting for confounders, two multivariable logistic regression models were applied. Model 1 analyzed all the sexually active women while model 2 analyzed sexually active women who had ever given birth only. In model 1, the factors that remained significantly associated with demand for LAPMs were: age of respondents, marital status, parity, rural/ urban residence, county of residence and access to FP information via media in the last 12 months (Table 3). Older sexually active women were less likely to have demand for LAPMs compared to adolescent women (15 – 19 years) [20 – 24 years: aOR = 0.31 (95% CI: 0.14, 0.68)]; 25 – 29 years: aOR = 0.19 (95% CI: 0.09, 0.42); 30 – 34 years: aOR = 0.15 (95% CI: 0.07, 0.34); 35 – 39 years: aOR = 0.18 (95% CI: 0.07, 0.43); 40 – 44 years: aOR = 0.24 (95% CI: 0.09, 0.62); 45 – 49 years: aOR = 0.16 (95% CI: 0.06, 0.39)]. Married women or women in union were 0.4 times less likely to have demand for LAPMs than unmarried sexually active women (aOR = 0.36; 95% CI: 0.23, 0.57). This study revealed that women who had 1–2 children were almost seven times more likely to have demand for LAPMs than women who had no children. There was a positive association between the demand for LAPMs and parity. The likelihood of having a demand for LAPMs increased with increase in the number of children the woman had [1 – 2 children: aOR = 7.16 (95% CI: 4.22, 12.13)]; 3 – 4 children: aOR = 28.04 (95% CI: 15.43, 50.95); 5+ children: aOR = 37.05 (95% CI: 18.40, 74.61)]. By residence, sexually active residing in rural setups were about 1.5 times more likely to have a demand for LAPMs than those who resided in urban setups (aOR = 1.29; 95% CI: 1.15, 1.45). Similarly, women who had accessed FP information via media in the last 12 months were about two times more likely to have demand for LAPMs than those who had not accessed (aOR = 1.51; 95% CI: 1.25, 1.82). This result showed a positive association between access to FP information via media and demand for LAPMs. By county of residence, while women from Kakamega county were less likely to have demand for LAPMs (aOR = 0.59; 95% CI: 0.38, 0.92) as compared to women from Bungoma county, women from Kericho, Kitui, Nandi and Siaya counties were more likely to have demand for LAPMs as compared to women from Bungoma county [Kericho: aOR = 1.92 (95% CI: 1.19, 3.10)]; Kitui: aOR = 1.42 (95% CI: 1.01, 2.10); Nandi: aOR = 1.56 (95% CI: 1.08, 2.26); Siaya: aOR = 1.74 (95% CI: 1.07, 2.82]. All significant covariates in model 1 were also significant in model 2 (which also had intendedness f the last pregnancy as an additional covariate) apart from access to FP information via media in the last 12 months.

**Table 3:** Multivariable analysis to identify respondents’ characteristics that may be associated with demand for LAPMs

|  | Model 1 |  | Model 2 |  |
| --- | --- | --- | --- | --- |
| Variable | Adjusted OR (95% CI) | P-value | Adjusted OR (95% CI) | P-value |
| Age |  |  |  |  |
| 15-19 | Ref. |  | Ref. |  |
| 20-24 | 0.31 (0.14, 0.68)** |  | 0.40 (0.16, 1.01) |  |
| 25-29 | 0.19 (0.09, 0.42)** |  | 0.26 (0.10, 0.65)** |  |
| 30-34 | 0.15 (0.07, 0.34)** | <0.001 | 0.20 (0.08, 0.51)** | <0.001 |
| 35-39 | 0.18 (0.07, 0.43)** |  | 0.24 (0.09, 0.63)** |  |
| 40-44 | 0.24 (0.09, 0.62)** |  | 0.31 (0.11, 0.90)* |  |
| 45-49 | 0.16 (0.06, 0.39)** |  | 0.23 (0.08, 0.67)** |  |
| Marital status |  |  |  |  |
| Never Married | Ref. |  | Ref. |  |
| Married/In a union | 0.36 (0.23, 0.57)** | <0.001 | 0.37 (0.20, 0.68)** | <0.001 |
| Widowed/Divorced | 0.69 (0.32, 1.49) |  | 0.75 (0.31, 1.82) |  |
| Parity |  |  |  |  |
| No children | Ref. |  |  |  |
| 1 or 2 children | 7.16 (4.22, 12.13)** | <0.001 | Ref. | <0.001 |
| 3 or 4 children | 28.04 (15.43, 50.95)** |  | 3.93 (3.02, 5.11)** |  |
| 5+ children | 37.05 (18.4, 74.61)** |  | 5.08 (3.35, 7.71)** |  |
| Residence |  |  |  |  |
| Urban | Ref. |  | Ref. |  |
| Rural | 0.72 (0.56, 0.93)* | 0.012 | 0.72 (0.55, 0.94)* | 0.018 |
| Education |  |  |  |  |
| Never | Ref. |  | Ref. |  |
| Primary/Technical | 1.13 (0.55, 2.33) |  | 1.11 (0.52, 2.40) |  |
| Secondary | 1.11 (0.52, 2.37) | 0.878 | 1.07 (0.48, 2.38) | 0.981 |
| Higher | 1.01 (0.46, 2.21) |  | 1.09 (0.47, 2.51) |  |
| Wealth |  |  |  |  |
| Lowest quintile | Ref. |  | Ref. |  |
| Lower quintile | 1.01 (0.73, 1.41) |  | 0.98 (0.70, 1.36) |  |
| Middle quintile | 1.17 (0.84, 1.62) | 0.733 | 1.15 (0.82, 1.61) | 0.833 |
| Higher quintile | 1.26 (0.85, 1.85) |  | 1.19 (0.79, 1.79) |  |
| Highest quintile | 1.26 (0.83, 1.91) |  | 1.16 (0.74, 1.8) |  |
| County |  |  |  |  |
| Bungoma | Ref. |  | Ref. |  |
| Kakamega | 0.59 (0.38, 0.92)* |  | 0.63 (0.39, 1.04) |  |
| Kericho | 1.92 (1.19, 3.1)** |  | 1.97 (1.18, 3.29)** |  |
| Kiambu | 0.83 (0.59, 1.19) |  | 0.90 (0.62, 1.30) |  |
| Kilifi | 0.72 (0.51, 1.02) |  | 0.75 (0.52, 1.09) |  |
| Kitui | 1.46 (1.01, 2.1)* | <0.001 | 1.45 (0.98, 2.13) | <0.001 |
| Nairobi | 0.75 (0.51, 1.11) |  | 0.80 (0.53, 1.20) |  |
| Nandi | 1.56 (1.08, 2.26)* |  | 1.54 (1.06, 2.23)* |  |
| Nyamira | 0.81 (0.56, 1.18) |  | 0.82 (0.56, 1.21) |  |
| Siaya | 1.74 (1.07, 2.82)* |  | 1.68 (1.02, 2.77)* |  |
| West Pokot | 0.51 (0.18, 1.44) |  | 0.51 (0.18, 1.43) |  |
| Access to FP information via media in the last 12 months |  |  |  |  |
| No | Ref. | **0.027** | Ref. | 0.079 |
| Yes | 1.53 (1.05, 2.23)* |  | 1.43 (0.96, 2.12) |  |
| Received FP information from health care workers in the last 12 months |  |  |  |  |
| No | Ref. | 0.522 | Ref. | 0.600 |
| Yes | 1.07 (0.88, 1.29) |  | 1.05 (0.87, 1.28) |  |
| Intendedness of the last pregnancy |  |  |  |  |
| Intended |  |  | Ref. |  |
| Unintended |  |  | 1.12 (0.89, 1.41) | 0.332 |
| Source of the current method |  |  |  |  |
| Private facility | Ref. | 0.205 | Ref. | 0.412 |
| Public facility | 1.17 (0.92, 1.49) |  | 1.11 (0.86, 1.43) |  |
| Told what to do if they experienced side effects |  |  |  |  |
| No | Ref. | 0.030 | Ref. | 0.045 |
| Yes | 0.69 (0.49, 0.96)* |  | 0.68 (0.46, 0.99)* |  |

Notes: Adjusted OR/ aOR – Adjusted Odds ratio; CI – Confidence interval

## DISCUSSION

The purpose of this study was to investigate the magnitude, recent trends and correlates of demand for LAPMs in Kenya. The study analyzed the PMA2020 data for the years 2014 to 2018 and included sexually active women of the reproductive age. The study did not include women who were pregnant at the time of the survey. In addition, women were not sure if they wanted a (another) child, women who did not know how long they would like to wait for their next birth and those women who had the time till next birth missing were also excluded as this information is required in the estimation of demand for LAPMs.(25)

As in this study, other studies in Kenya have found out that there has been a general shift towards the use of long-acting reversible methods especially implants.(37) This study results demonstrate a steady increase in the utilization of modern contraceptives and particularly the LAPMs among sexually active women in Kenya. On the other hand, there is a decrease in the use of short-acting methods of contraception. These findings are similar to those reported by others.(15)(38) Tsui et. al. (2017) in their study assessing the modern contraceptive practice levels and trends in SSA and other world regions observed that the highest levels of modern method use among married women to be in countries in the Southern and Eastern African regions, and they noted that contraceptive uptake have increased more quickly in Eastern relative to Western Africa.(39) Similarly, according to Cahill et. al. (2018) Mozambique and Kenya had the largest positive changes between 2012 and 2017, with an increase in mCPR of 15·7% (95% UI 4·6–23·0) in Mozambique and 12·7% (2·3–22·5) in Kenya over the 5 years.(40) In Kenya the increase in LAPMs uptake has largely been driven by implant guarantee and increased efforts by Kenya government to improve access to LAPM as well as by the increase in the use of implants, which has steadily overtaken uptake of pills, but still lagging behind injectables.(38) This was evident in this study as we observed that injectables were the preferred method among sexually active contraceptives users who desired to have a child then or within 2 years, among contraceptives users who desired to delay or avoid pregnancy for 2 years or more and among women who did not want any more children. This was also the case in other studies in SSA.(39)

In addition even though there has been an increase in share of LAPMs use, a large proportion of those who either want to space their births for two or more years or who want to limit their family sizes are still using methods which are unlikely to meet their desires. That is nearly a third (30.2%) of the sexually active women desired to delay or avoid pregnancy for 2 years or more and 30.8% of women would want to stop childbearing but continue to depend on short acting methods. The results portrayed here may be consistent with similar observations made in Ghana where educated urban women achieve desired family goals using a blend less effective methods such as abstinence, withdrawal, condoms or emergency contraception (Marston et al., 2017).

The study also found that the demand for LAPMs as well the total unmet need for LAPMs among sexually active women declined substantially between 2014 and 2018. These results were consistent with other studies. Cahill et. al. (2018) found that unmet need for contraception decreased between 2012 and 2017 with Kenya making the most progress and decreasing unmet need by 7·8 percentage points (95% UI 1·1 to 14·1) in 5 years, followed by Malawi with a drop of 6·1 percentage points (− 0·4 to 12·6) over the same period.(40) The substantial decrease in the unmet need for LAPMs suggests that FP investments are keeping pace with the desires of married women to prevent pregnancy. Decrease in the demand for LAPMs implies that great efforts would be needed both to generate demand and to address existing demand.

In this study age of respondents, marital status, parity, rural/ urban residence, county of residence and access to FP information via media in the last 12 months were significantly associated with demand for LAPMs. From our findings, women age 15-19 years had higher odds of having a demand for LAPMs than other women. This result in line with the result of a study from five countries using reports of over 100 active surveillance sites where women at greatest risk for having an unmet need (a component of demand) for family planning were young women below the age of 20 years.(41) In this study, marital status and parity were also identified as some of the factors determining demand for LAPMs. The likelihood of having a demand for LAPMs among sexually active women increased with increase in the number of children the woman had. It can be argued that this may be necessitated by the fact that the more child the woman is having, the more likely she may want to space or limit the number of child and the more she may need a more effective method of contraception. These findings are in sync with findings from other studies where marital status and parity have been identified as important individual characteristics influencing women’s reproductive health behaviors, including demand and uptake of modern contraception.(42–44) In addition, we observed that respondents who had access to FP information via media in the last 12 had higher odds of having a demand for LAPMs. This result is supported by studies done elsewhere.(45,46) There was also lower odds of demand for LAPMs among rural residents as compared to urban residents. This is in line with a study done in Ethiopia which found that being an urban resident was positively associated with demand for long acting contraceptive methods.(47) This can be attributed to the fact that access to family planning services especially the LAPMs in sub-Saharan Africa is especially low among rural.(48)

## Conclusion

Though Kenya is among the leading countries in contraceptives utilization in SSA, the uses of long-acting methods like IUCD and permanent methods were seen to be very low. In this study we found that the total demand for LAPMs among sexually active women in Kenya is high, with the bigger share of the demand being unmet. Woman’s age, marital status, parity, rural/ urban residence and county of residence were significantly associated with demand for LAPMs among this group of women. Therefore, targeted programs to address existing demand should be put in place to ensure no woman is left behind. In addition, access to FP information via media in the last 12 months was also significantly associated with demand for LAPMs. Therefore programmers and advocates should utilize the locally available media to generate demand.

## Data Availability

PMA2020 data are accessible on request through the project website upon approval by PMA2020's coordinating center at JHU in Baltimore. However, data used for this analysis can be made available by the research team to researchers who meet the criteria for access to data

https://www.pma2020.org/request-access-to-datasets-new

## CONTRIBUTORS

MWW conceived the ideas of this analysis, developed the statistical analysis plan, analysed and interpreted the data. PG was Principal Investigator of the PMA study, contributed in the conception and design of the study, and also in the data acquisition. MT trained and supervised data collectors. MWW led the manuscript writing with substantive input from PG. All authors reviewed, edited drafts and approved the final version.

## ACKNOWLEDGEMENTS

We appreciate the support of PMA/Kenya central staff and the pool of female data collectors. The data collection funding was provided by the Bill & Melinda Gates Foundation. No additional funding was sought to complete this article. The funding body had no role in the design of the study, data collection, analysis, and interpretation of data and in writing the manuscript. The manuscript represents the view of the named authors only.

